# Diagnostic accuracy of the Cobas^®^ MTB and Cobas^®^ MTB/RIF-INH assays on sputum and the Cobas^®^ MTB assay on tongue swabs for *Mycobacterium tuberculosis* complex detection in symptomatic adults in South Africa

**DOI:** 10.1101/2025.06.30.25330606

**Authors:** Anura David, Lyndel Singh, Pedro da Silva, Keneilwe Peloakgosi-Shikwambani, Zanele Nsingwane, Violet Molepo, Wendy Stevens, Lesley Scott

**Affiliations:** Wits Diagnostics Innovation Hub, Health Sciences Research Office, Faculty of Health Sciences, University of the Witwatersrand, Johannesburg, South Africa; National Priority Programmes, National Health Laboratory Services, Johannesburg, South Africa

**Author notes:** Corresponding author: Anura David.

## Abstract

Accurate and rapid detection of *Mycobacterium tuberculosis* complex (MTBC) and drug resistance is essential for effective tuberculosis (TB) management, particularly in high-burden settings. The Cobas® MTB and Cobas MTB/RIF-INH assays are moderate-complexity nucleic acid amplification tests that detect MTBC and resistance to rifampicin (RIF) and isoniazid (INH).

This study evaluated clinical diagnostic performance of the Cobas assays on sputum, using liquid culture as the reference standard and Xpert MTB/RIF Ultra (Xpert Ultra) for comparison. Diagnostic accuracy of the Cobas MTB assay on tongue swabs (TS) was also assessed.

In a study population (n=354) with 56% HIV prevalence, the overall sensitivity and specificity of Cobas MTB on sputum was 93.8% (95% CI: 84.8-98.3) and 100% (95% CI: 98.7-100) compared with culture. The assay showed almost perfect agreement with Xpert Ultra (Cohen’s kappa = 0.904). Among HIV-positive participants, sensitivity was 88.2% (95% CI: 97.8-100). RIF resistance profiling by Cobas MTB/RIF-INH was fully concordant with culture and Xpert Ultra. Three INH-resistant cases were missed, likely due to genotypic-phenotypic discordance. Although specimen numbers were small, TS demonstrated better diagnostic accuracy when using a diluted (66%) Microbial Inactivation Solution.

The Cobas MTB and MTB/RIF-INH assays demonstrated high diagnostic accuracy compared to culture and Xpert Ultra on sputum. Findings support TS as an alternative specimen type for MTBC detection using an optimized protocol. These findings underscore the potential of the Cobas assays as reliable alternatives for TB and resistance diagnostics, particularly in settings where rapid, accurate detection of MTBC and RIF or INH resistance is crucial.

**Importance:** The study demonstrates high diagnostic accuracy of the Cobas® MTB and MTB/RIF-INH assays, on sputum, when compared with culture and Xpert Ultra while offering critical advantages, namely, INH resistance detection and scalability via high-throughput platforms. Preliminary findings on tongue swabs indicate potential for additional specimen types with modified pre-processing protocols. This research supports the incorporation of the Cobas assays into diversified TB testing strategies to improve detection and management in high-burden settings.

## Introduction

The Cobas® 6800/8800 Systems (Roche Molecular Systems Inc., Pleasanton, CA) have been integral to South Africa’s diagnostic landscape. These high-throughput platforms were introduced in 2015 (1) for testing of the Cobas® HIV-1 quantitative nucleic acid amplification test (NAAT), and has since played a critical role in HIV management in a country with one of the largest populations of people living with HIV (PLHIV). The Cobas systems have additionally, demonstrated versatility by supporting alternative specimen types, such as plasma separation cards (2), thereby providing an option to testing conventional plasma samples.

It is not surprising that a country with such high HIV rates also ranks as one of the countries with the highest burden of tuberculosis (TB) and multi-drug resistant TB (MDR-TB), globally (3). For TB diagnosis, the Xpert MTB/RIF (Xpert) assay was used as the initial diagnostic since 2011, followed by the Xpert MTB/RIF Ultra (Xpert Ultra) assay in 2017. Shortage of Xpert Ultra cartridges experienced during (and after) the COVID-19 pandemic highlighted the risks with reliance on a single supplier. This shortage together with the growing diagnostic pipeline (4) as well as the WHO recommendation for use of a new category of diagnostics, the moderate complexity assays in 2021 (5), led to the diversification of the molecular platforms used in the South African National TB program (6). Since 2023, alongside the Xpert Ultra assay, the Cobas MTB and MTB RIF/INH assays (Roche Molecular Systems Inc., Pleasanton, CA) and the MAX MDR-TB (Becton, Dickinson and Company, Sparks, MD, USA) assays have been incorporated into the TB testing algorithm. Since the Cobas systems were already in use for the HIV program with established in-country support, integrating a test utilizing the same technology provided a strategic advantage.

The Cobas MTB assay is an automated, qualitative, real-time PCR test designed to detect *Mycobacterium tuberculosis* complex (MTBC) DNA in respiratory specimens. If a MTBC-positive result is detected on the assay, the specimen can be reflexed to the Cobas MTB/RIF-INH assay which detects rifampicin (RIF)-resistance associated mutations of the *rpoB* gene, and isoniazid (INH)-resistance associated mutations in the *katG* and *inhA* genes. Both assays’ were previously evaluated by our group, first in an analytical evaluation using spiked sputum specimens and culture isolates (7) and then in a multi-country clinical performance evaluation (8). However, the clinical evaluation study design excluded individuals unable to produce ≥2 mL of sputum, effectively omitting those who typically present for TB investigation in SA with sputum volumes around 1 mL. We previously evaluated the Cobas MTB assay (9) but the comparator used in that study was Xpert, not the currently used Xpert Ultra.

Other studies have assessed performance of the Cobas TB assays using N-acetyl-l-cysteine-sodium hydroxide (NALC-NaOH)-treated specimens (10) or culture isolates and spiked sputum (11) but data on raw sputum is limited.

In this clinical performance evaluation on sputum, we assessed the performance of the Cobas MTB and Cobas MTB-RIF/INH assays for the detection of MTBC, RIF and INH resistance compared to a liquid culture reference standard and to Xpert Ultra as a comparator. Given our group’s focus on exploring tongue swabs (TS) as an additional specimen type for TB diagnosis, we collected TS in parallel with sputum and evaluated the diagnostic accuracy of the Cobas MTB assay for MTBC detection in TS specimens.

## Materials and Methods

### Study Design and procedures

We performed a cross sectional, prospective diagnostic study in which the accuracy of the Cobas MTB assay performed on sputum was assessed using the Mycobacterial Growth Indicator Tube (MGIT) (Becton Dickinson, Sparks, MD, USA) liquid culture reference standard. Cobas MTB assay performance was additionally compared to that of the Xpert Ultra assay. Additionally, concordance of the Cobas MTB assay on tongue swabs (TS) was evaluated using Xpert Ultra, liquid culture, and Cobas MTB results from sputum as comparators.

Symptomatic adults (≥18 years) attending the Hillbrow Community Health Centre (HCHC), in Johannesburg, Gauteng, South Africa, and being investigated for TB, were approached and invited to enroll in the study. Recruitment occurred from 20 October 2021 to 11 July 2023. The research nurse performed the WHO-recommended four-symptom screen (W4SS)—cough, fever, weight loss, and night sweats. Personal characteristics such as age, weight, height, HIV status and previous TB history were also recorded. Weight and height measurements were used to calculate Body Mass Index (BMI) which was used as an indicator of nutritional status. Participants with a BMI below 18.5 kg/m² were classified as underweight, suggesting possible malnutrition. Study inclusion criteria included participant willingness to return for a second visit, provision of the required number of study specimens and absence of any TB treatment within six months prior to enrolment. Specimen collection was performed for routine and research testing, over two visits (Figure 1). Research nurses used two Copan FLOQSwab (Copan, Brescia, Italy) swabs for tongue collection from each participant using the procedure detailed by Andama *et al.* (12). Collection was performed by swabbing the dorsum of the tongue for 30 seconds, as far back on the tongue as possible without initiating a gag reflex.

**Figure 1:**
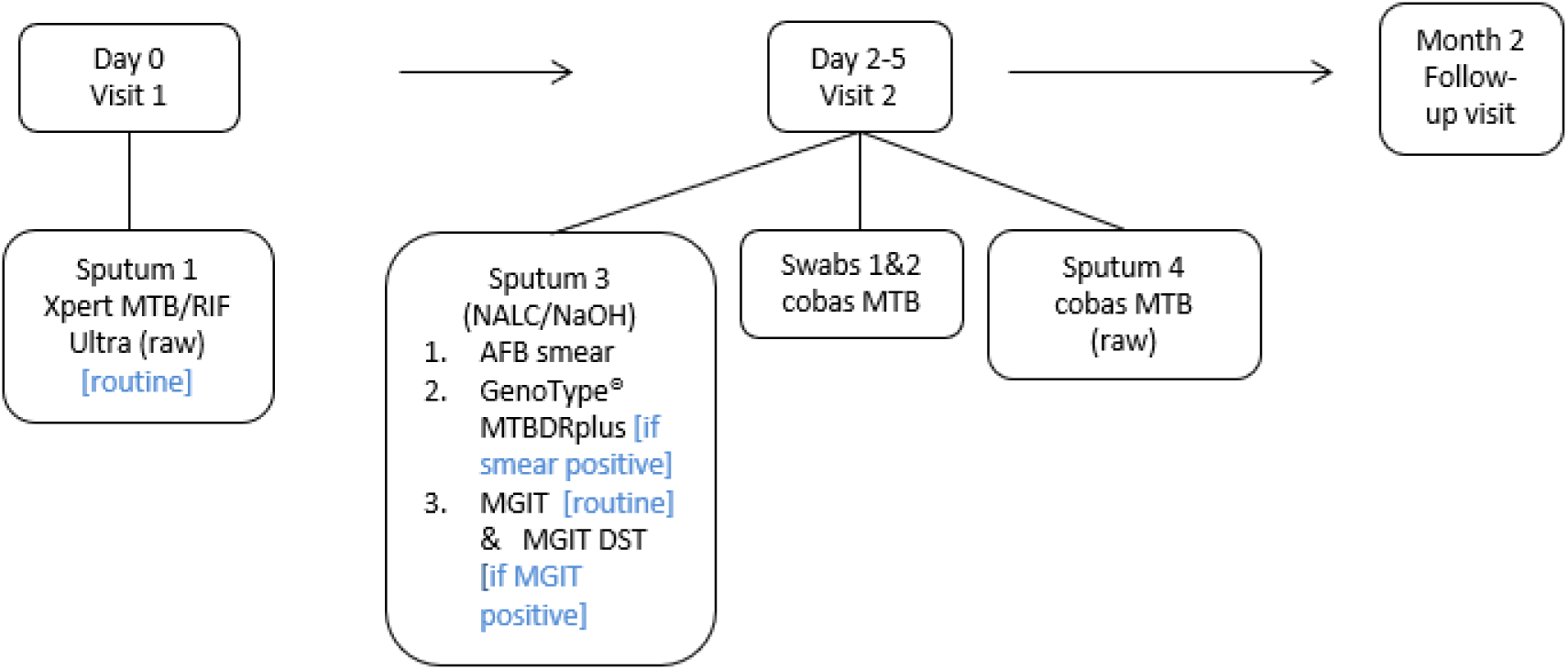
Description of study outline indicating clinic visits and specimen laboratory pathways Specimens for routine and study testing were collected over two visits approximately 2-5 days apart. AFB, acid-fast bacilli; MGIT, Mycobacterial Growth Indicator tube; DST, drug-susceptibility testing; NALC/NaOH,N-acetyl-L-cysteine–sodium citrate–sodium hydroxide; BD, Becton Dickinson

One TS was collected before sputum collection and the second after sputum collection. Participants abstained from consuming any food or beverages for at least 30 minutes prior to specimen collection. Both swabs were transported “dry” (without any transport buffer) to the laboratory for testing.

Routine and research testing was performed at the Wits Diagnostic Innovation Hub (WitsDIH) research laboratory in Braamfontein (Johannesburg).

### Ethics statement

Ethics approval for this study was obtained from the University of the Witwatersrand Human Research Ethics Committee (M1911150). The trial was registered with the South African National Clinical Trials Registry (DOH-27-052021-5442). All participants provided written informed consent.

### Laboratory testing

Genotype® MTBDRplus line probe assay (LPA) (Hain Lifescience/Bruker, Nehren, Germany) testing was performed on all smear-positive sputum and also used to speciate liquid cultures that were acid-fast bacilli (AFB) positive. Routine testing and result reporting was performed by laboratory staff in accordance with the National Tuberculosis Management Guidelines (13). Staff performing routine testing were blinded to Cobas MTB results.

Cobas MTB testing on sputum was performed according to manufacturer instructions. Sputum was stored at −20°C until batch testing. Microbial Inactivation Solution (MIS) (Roche Molecular Systems Inc., Pleasanton, CA) was added to raw sputum in a 2:1 ratio of solution to specimen. This mix was incubated at room temperature for 60 minutes, followed by sonication and a 5-minute centrifugation. Testing was performed on the Cobas 6800 System. Results are reported as MTB Positive, MTB Negative or Invalid (test was unable to produce a reliable or interpretable result due to issues with the specimen or the testing process). Any specimen which produced an MTB Positive result, was reflexed for RIF and INH testing using the Cobas MTB/RIF-INH assay. Research staff performed Cobas testing and were blinded to routine TB results.

Swabs were stored at −80°C until batch testing. For processing of TS, 1.8 mL of MIS was added to the “dry” swab and 1.5 mL was loaded on the Cobas 6800 System. Processing of TS was performed according to the standard sputum protocol using neat MIS, with the exception that the centrifugation step was excluded (n=239). However, parallel testing on contrived swabs showed improved sensitivity using a diluted (66%) MIS buffer, hence 66% MIS buffer was used to process the remaining swabs (n=99).

### Outcomes and statistical analysis

For diagnostic accuracy performance, sputum results from the Cobas MTB assay were compared with liquid culture for MTBC detection and to phenotypic drug susceptibility testing (pDST) (performed using the MGIT960 SIRE kit (Becton Dickinson, Sparks, MD, USA)) for RIF and INH resistance detection. Data analysis included calculation of sensitivity, specificity, positive predictive value (PPV) and negative predictive value (NPV) for MTBC detection and concordance for RIF and INH resistance detection, with 95% CIs calculated using the Wilson score method (first including all participants and then excluding participants with an Ultra “trace” result). Cobas MTB results were additionally compared with Xpert Ultra. For Cobas MTB assay performance on TS, concordance with Xpert Ultra, liquid culture and Cobas MTB sputum is reported. The target sample size was 400 participants which was derived using the formula described below with population proportion estimated at 50% and 95% confidence interval set at 5%. Sample size (n) = (Z)^2*p(1-p) / x^2, Where Z = 1.96, & where p = population proportion & where x = confidence interval as a proportion. In determining the sample size, an allowance was made for participants who do not meet the inclusion criteria. To determine the performance of the Cobas MTB assay, only specimens that generated valid results across all tests (Cobas MTB, Ultra and MGIT) were included in the statistical analysis.

## Results

### Characteristics of the study population

A total of 554 participants were screened for the study (Figure 2). Of these, 133 were excluded during the recruitment process, and 421 participants were consented and enrolled. Of those enrolled, 67 were excluded from the analysis, resulting in a final sample size of 354 participants included in the statistical analysis. Characteristics of the study population are shown in Table 1. The average age of participants was 39 years and most (64%) were male. As indicated by their BMI, a total of 28/66 (42%) participants diagnosed with active TB were malnourished. Among the 351 participants with a known HIV status, 57% (199/351) were positive. Forty-two reported a previous TB episode. Tuberculosis was microbiologically confirmed on liquid culture in 64/354 (18%) of participants. A total of 290 participants did not have microbiologically confirmed tuberculosis (TB) and were consequently classified as not having active TB disease. The culture contamination rate was 11% (43/404); of these, the Ultra and Cobas MTB assays detected one MTBC-positive specimen, and the participant reported an improvement in symptoms after having received treatment. Sixty-six participants were microbiologically confirmed to have TB using the Xpert Ultra assay.

**Figure 2:**
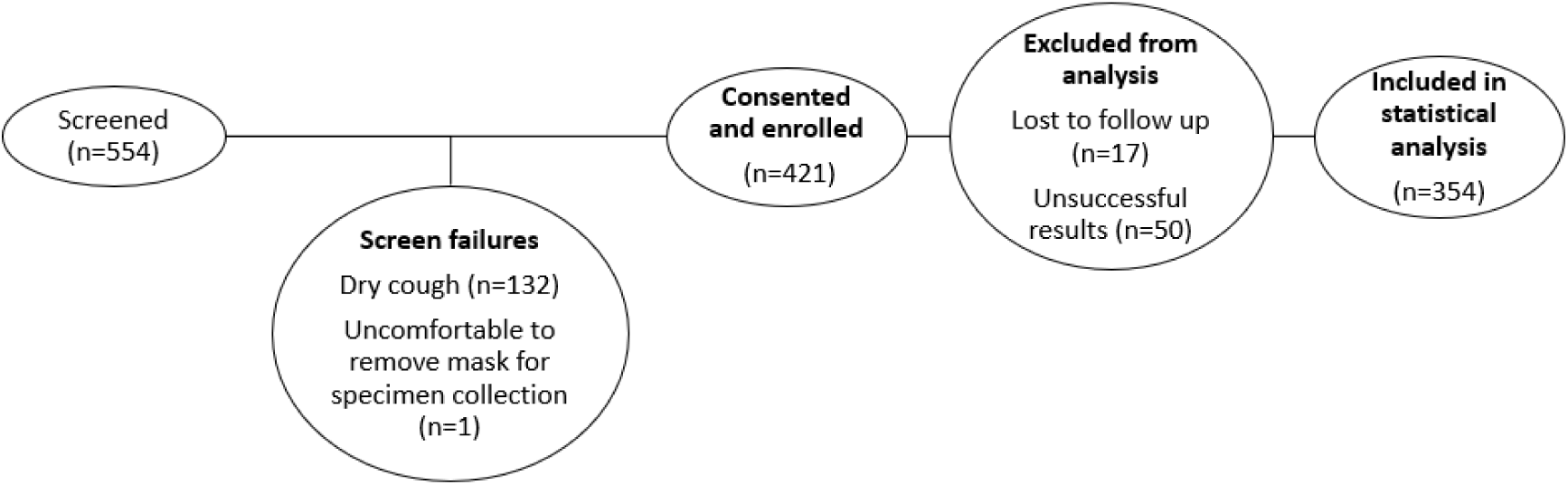
Data description for statistical analysis Of 554 individuals screened, 133 were excluded (132 with dry cough and 1 unwilling to remove a mask for specimen collection). A total of 421 participants were enrolled and consented. Of these, 67 were excluded from analysis due to loss to follow-up (n = 17) or unsuccessful test results (n = 50). The final dataset included 354 participants for statistical analysis.

**Table 1:** Cohort characteristics for participants used in the statistical analysis.

| Characteristic | All (n=354) |
| --- | --- |
| *Demographics* |  |
| Age, mean (range), years | 39 (18-70) |
| Male sex, n (%) | 213 (63.6) |
| BMI kg/m<sup>2</sup>, n (%) |  |
| <18.5 (Underweight) | 94 (26.6) |
| 18.5-24.9 (Healthy weight) | 186 (52.5) |
| 25-29.9 (Overweight) | 44 (12.4) |
| >30 (Obese) | 30 (8.5) |
| *HIV-related information* |  |
| HIV-positive, n (%) | 199 (56.2) |
| HIV-negative, n (%) | 152 (42.9) |
| Status unknown, n (%) | 3 (0.9) |
| *TB History* |  |
| Previously diagnosed with TB, n (%) | 42 (11.9) |
| *Clinical signs and symptoms of TB at presentation* |  |
| Cough (any duration), n (%) | 353 (99.7) |
| Unexplained weight loss, n (%) | 265 (74.9) |
| Nights sweats, n (%) | 263 (74.2) |
| Fever, n (%) | 236 (66.9) |
| *Other, n (%) | 257 (72.6) |
| *Bacteriological confirmation (sputum 3), n (%)* |  |
| Smear and culture positive | 44 (12.4) |
| Smear negative and culture positive | 20 (5.6) |
| Smear and culture negative | 284 (80.2) |
| Smear positive and culture negative | 6 (1.7) |
| Xpert MTB/RIF Ultra (sputum 1) | 66 |
| High | 24 |
| Medium | 8 |
| Low | 19 |
| Very low | 8 |
| Trace | 7 |

BMI, body-mass index; \*Other symptoms include body aches, loss of appetite, hemoptysis, shortness of breath, dizziness and vomiting

### Clinical performance evaluation of the Cobas MTB assay on sputum

The performance of the Cobas MTB assay on raw sputum was compared with the reference method of liquid culture and, additionally, stratified by HIV and smear status, as outlined in Table 2 for 354 specimen results. The assay exhibited high sensitivity (94%) and 100% specificity, with improved performance observed when Xpert Ultra “trace” results were excluded. Assay sensitivity among HIV-positive individuals was 88%, while perfect sensitivity (100%) and specificity (100%) was achieved in HIV-negative individuals. The Cobas MTB assay produced 1/404 (<1%) invalid results. Unsuccessful tests were not repeated due to cost considerations.

**Table 2:**
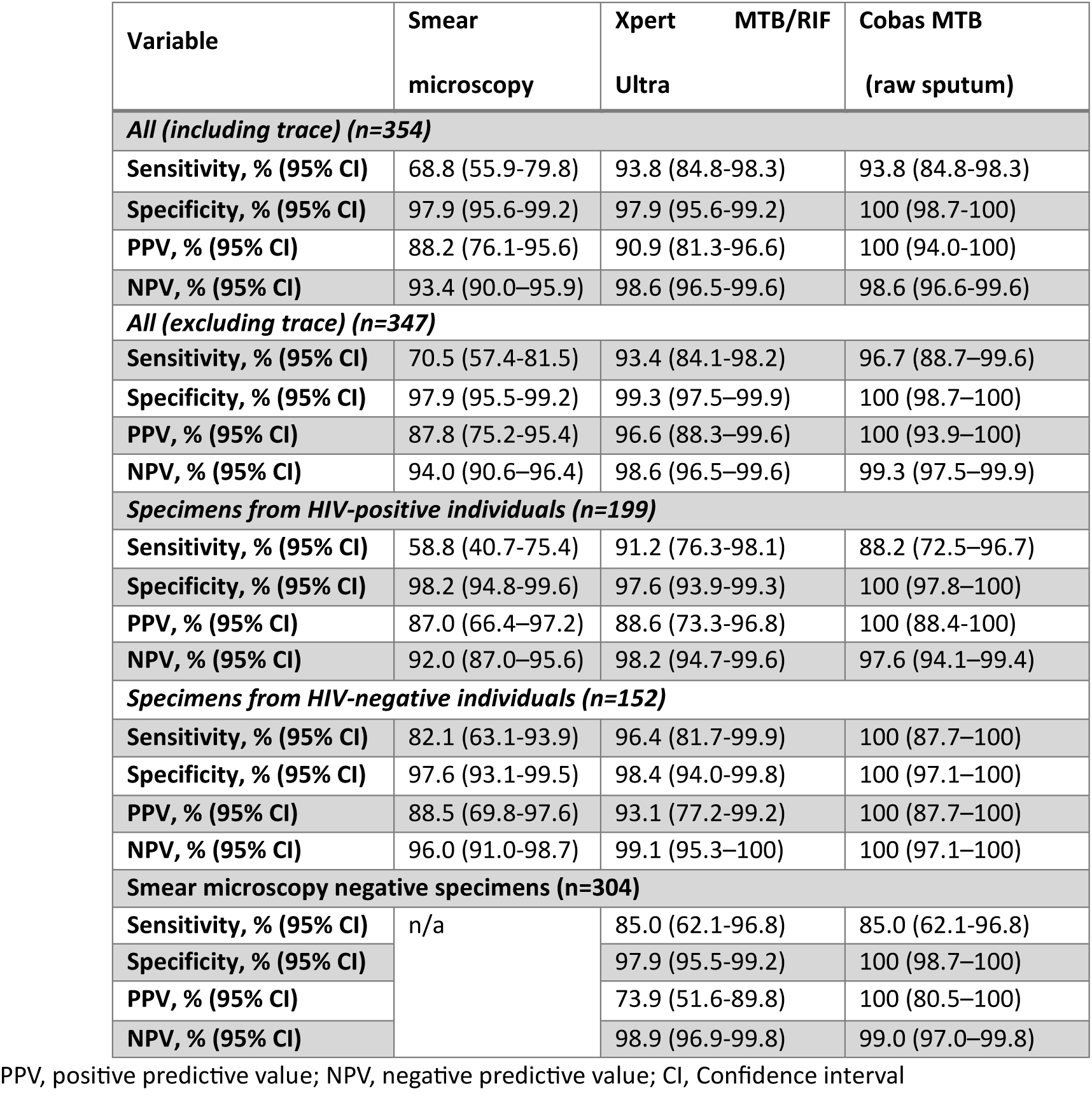
The performance of smear microscopy, Xpert Ultra and Cobas MTB assays compared to the reference standard (liquid culture) for MTBC detection on sputum.

### Comparison of the Cobas MTB assay to Xpert Ultra for Detection of MTBC

The Cobas MTB and Xpert Ultra assays demonstrated a high level of concordance, with an overall agreement of 97.5% (344/354) and a Cohen’s kappa coefficient of 0.904 (95% CI: 0.8447–0.9623).

Among the discordant results, eight specimens were positive by Xpert Ultra but negative by Cobas MTB; all eight had “trace” or “very low” semi-quantitative results on Ultra, and two of these were culture-positive. Conversely, two specimens were positive by Cobas MTB but negative by Xpert Ultra; both were also confirmed positive by liquid culture.

### Clinical performance evaluation of the Cobas MTB/RIF-INH assay on sputum

Of the 60 raw sputum which produced an MTB Positive result on the Cobas MTB assay and were reflex tested on the Cobas MTB/RIF-INH assay, reportable resistance results were available for 52 (87%) specimens. Unsuccessful tests were not repeated due to insufficient specimen volumes.

For RIF resistance detection, the Cobas MTB/RIF-INH assay accurately identified correct resistance profiles in 47/47 (100%) sputum samples with valid results (Table 3). Of the specimens tested, 8/60 (13%) yielded invalid results. Where pDST was unsuccessful, the assay successfully assigned resistance profiles to five specimens, as confirmed by the LPA.

**Table 3:** Cobas RIF resistance results, compared to phenotypic DST.

| pDST result | Cobas MTB-RIF/INH result |  |  |
| --- | --- | --- | --- |
|  | RIF-R<br>not detected | RIF-R<br>detected | Invalid |
| **RIF sensitive (n=54)** | 46 | 0 | 8 |
| **RIF resistant (n=1)** | 0 | 1 | 0 |
| **Invalid (n=5)** | 5 | 0 | 0 |

RIF, rifampicin; RIF-R, rifampicin resistance; pDST, phenotypic drug susceptibility testing

For INH resistance detection, the Cobas MTB/RIF-INH assay correctly identified resistance profiles in 44/47 (94%) sputum samples with valid results (Table 4). In three cases, the assay did not detect INH resistance that was identified by pDST, with resistance also undetected by LPA in 1/3 (33%) of these cases. Additionally, 8/60 (13%) specimens produced invalid results; however, the assay successfully assigned resistance profiles to five specimens, as confirmed by LPA, where pDST was unsuccessful.

**Table 4:** Cobas INH resistance results, compared to phenotypic DST.

| pDST result | Cobas MTB-RIF/INH result |  |  |
| --- | --- | --- | --- |
|  | INH-R<br>not detected | INH-R<br>detected | Invalid |
| INH sensitive (n=50) | 43 | 0 | 7 |
| INH resistant (n=5) | 3 | 1 | 1 |
| Invalid (n=5) | 5 | 0 | 0 |

INH, isoniazid; RIF-R, isoniazid resistance; pDST, phenotypic drug susceptibility testing

### Diagnostic performance of tongue swabs on the Cobas MTB assay

When compared to Xpert Ultra, TS detected MTBC across all semi-quantitative categories, from “high” to “very low.” Using undiluted MIS, MTBC detection was observed in three participants on TS collected after sputum, compared to those collected before (Table 5). However, with 66% MIS, detection improved across all semi-quantitative categories, with comparable detection rates between TS collected before and after sputum. Similarly, when compared to liquid culture or Cobas MTB sputum and processed with 66% MIS, TS showed improved MTBC detection and comparable detection rates on Cobas MTB. Regardless of the comparator used, assay specificity remained consistent between undiluted and 66% MIS.

**Table 5:** Performance of the MAX MDR-TB assay on tongue swabs compared to Xpert Ultra, liquid culture and MAX MDR-TB sputum results.

|  | Cobas MTB TS result, n/N (%) |  |  |  |
| --- | --- | --- | --- | --- |
| Comparator test<br>result (sputum) | Neat MIS<br>(TS 1) | Neat MIS<br>(TS 2) | 66% MIS<br>(TS 1) | 66% MIS<br>(TS 2) |
| *Xpert Ultra positive* | 25/53 (47.2) | 28/53 (52.8) | 9/13 (69.2) | 9/13 (69.2) |
| *High* | 16/18 (88.9) | 15/18 (83.3) | 4/6 (66.7) | 4/6 (66.7) |
| *Medium* | 2/6 (33.3) | 3/6 (50.0) | 1/2 (50.0) | 1/2 (50.0) |
| *Low* | 6/15 (40.0) | 6/15 (40.0) | 4/4 (100) | 4/4 (100) |
| *Very low* | 1/7 (14.2) | 4/7 (57.1) | 0/1 (0) | 0/1 (0) |
| *Trace* | 0/7 (0) | 0/6 (0) | - | - |
| **Xpert Ultra**<br>**negative** | 215/217 (99.1) | 216/217<br>(99.5) | 70/71 (98.6) | 71/71 (100) |
| **Liquid culture**<br>**positive** | 26/52 (50) | 29/52 (55.8) | 9/12 (75.0) | 9/12 (75.0) |
| **Liquid culture**<br>**negative** | 217/218 (99.5) | 218/218<br>(100) | 71/72 (98.6) | 72/72 (100) |
| **Cobas MTB sputum**<br>**positive** | 26/48 (54.2) | 29/48 (60.4) | 9/12 (75.0) | 9/12 (75.0) |
| **Cobas sputum**<br>**negative** | 221/222 (99.5) | 222/222<br>(100) | 71/72 (98.6) | 72/72 (100) |

MIS, microbial inactivation solution; TS, tongue swab; TS 1 was collected before sputum collection and TS 2 after sputum collection

## Discussion

This study investigated the clinical performance of the Cobas MTB and Cobas MTB/RIF-INH assays for MTBC, RIF and INH resistance detection using raw sputum against a liquid culture reference standard and to Xpert Ultra as a comparator. In addition, we investigated the diagnostic accuracy of the Cobas MTB assay on TS compared with Xpert Ultra sputum, liquid culture and Cobas MTB sputum.

In this study population with an HIV prevalence of 57%, the Cobas MTB assay demonstrated high sensitivity (94%) compared to culture, comparable to the Xpert Ultra assay (94%), with a specificity of 100%. Among HIV-positive participants, the Cobas MTB assay showed a sensitivity of 88%, compared to 91% for Xpert Ultra. In this subset, when participants with “trace” semi-quantitative Xpert Ultra results were excluded from the analysis, the sensitivity of the Cobas MTB assay improved to 97%. The assay exhibited perfect sensitivity (100%) within the HIV-negative subset of participants. There was almost perfect agreement (14) between the Cobas MTB and Xpert Ultra assays, reflecting a high degree of concordance with both assays demonstrating similar unsuccessful rates of <1%. Thus, in terms of performance, both assays perform comparably which supports use of the Cobas MTB assay in South Africa’s TB testing diversification.

Xpert Ultra has the advantage of providing a MTBC result and an upfront RIF resistance profile within a shorter period and fewer pre-processing steps compared to the Cobas MTB assay (15). However, the Cobas assay, despite requiring reflex resistance testing can provide an INH result which the Xpert Ultra cannot. This has implications for patient management, as a South African prevalence survey (2012–2014) found that INH-monoresistant TB accounted for more than 5% of cases in all provinces (16). Since testing is performed on the high throughput 68/8800 systems, the Cobas MTB assay is suited for high-burden settings or in countries which are trying to ramp up TB screening in attempts to overcome the effects of COVID-19 (17). An additional advantage of the Cobas MTB assay is that resistance testing can be performed only when required, which will save costs.

For RIF resistance detection, the Cobas MTB/RIF-INH assay showed perfect agreement with liquid culture and Xpert Ultra. Both Xpert Ultra and Cobas MTB/RIF-INH yielded an unsuccessful rate of 13% for RIF detection. For INH, resistance was missed in three specimens (as per the pDST result); one of which was also missed by the LPA. One explanation for this is that it has been shown that phenotypic resistance may sometimes be identified before the associated genetic mutations can be detected since molecular assays require a higher drug resistant proportion to be present. This mismatch can arise from the limited sensitivity of genotypic assays, which generally focus on the most frequently occurring mutations, or from resistance mechanisms that have not yet been genetically characterized (18).

For the Cobas MTB assay on TS, MTBC detection was comparable between TS collected before and after sputum collection, suggesting that the timing of collection does not affect assay performance. When evaluated against Xpert Ultra sputum semi-quantitative results, the Cobas MTB assay on TS detected MTBC down to the “very low” category, with improved detection in participants with higher bacterial loads, though some variability was observed. These findings align with those reported by Ahls et al. (19), further supporting that MTBC detection using TS improves with increasing bacterial load. Cobas MTB assay performance on TS is lower than that on sputum but this is expected since the assay is designed for sputum and additional specimen types will likely require protocol modification, as has been seen with other assays designed for sputum but tested on TS (20). Some protocol modification was attempted in this study, where a diluted MIS (66% MIS) was tested on a subset of specimens to determine if assay sensitivity could be improved. While the number of MTBC positive specimens is too limited to draw definitive conclusions, assay sensitivity and specificity seems improved using 66% MIS. Future studies should consider this diluted buffer for TS pre-processing for testing on the Cobas MTB assay.

In conclusion, the Cobas MTB and Cobas MTB/RIF-INH assays demonstrated comparable performance to liquid culture and the Xpert Ultra assay, on sputum, showcasing high sensitivity and specificity in detecting MTBC. These findings underscore the potential of the Cobas assays as reliable alternatives for TB and resistance diagnostics, particularly in settings where rapid, accurate detection of MTBC and RIF or INH resistance is crucial. The study findings further support the potential utility of TS as an additional specimen type for MTBC detection on the Cobas MTB assay, particularly when using an optimized pre-processing protocol such as 66% MIS. Future studies should explore application of the Cobas assays in diverse patient populations and operational settings to further establish utility in global TB control efforts.

## Data Availability

All data produced in the present study are available upon reasonable request to the authors

## Acknowledgments

We thank the study participants; the Johannesburg Health District and the and the Gauteng Department of Health for their support and collaboration on this study; Roche Molecular Systems Inc. for providing technical support, kits and equipment for this study. Roche Molecular Systems Inc. was not involved in the study design and analysis and interpretation of results.

## Author contributions

Conceptualization: A. David and L. Scott; Data curation: A. David; Formal analysis: A. David; Funding acquisition: L. Scott and W. Stevens; Investigation: A. David, L. Singh. K. Peloakgosi-Shikwambani, Z. Nsingwane and V. Molepo; Methodology: A. David and L. Scott; Project administration: L. Scott and W. Stevens; Supervision: L. Scott; Validation: A. David; Visualization: A. David and L. Scott; Writing—original draft: A. David; Writing—review and editing: A. David, L. Scott, P. da Silva, L. Singh, K. Peloakgosi-Shikwambani, Z. Nsingwane. V. Molepo and W. Stevens.

